# Time-trends in life expectancy of people with severe mental illness in Scotland, 2000-2019: a population-based study

**DOI:** 10.1101/2024.07.25.24310972

**Authors:** Kelly Fleetwood, Raied Alotaibi, Stine H Scheuer, Daniel J Smith, Sarah H Wild, Caroline A Jackson

## Abstract

**Objective:** To determine time-trends in life expectancy (LE) of people with a severe mental illness (SMI) compared with the general population.

**Design:** Observational population-based study.

**Setting:** Scotland, 2000-2019. Linked psychiatric hospital admission and death records.

**Participants:** Adults with a psychiatric hospital admission record for schizophrenia (28,797), bipolar disorder (16,657) or major depression (72,504) compared with the Scottish population (4.3 million adults in 2011).

**Main outcome measures:** Trends over time in life years lost for people with schizophrenia, bipolar disorder or major depression compared with the Scottish population, for all deaths, and natural and unnatural deaths, stratified by sex.

**Results:** Among people with SMI, one third died during the study period. Between 2000 and 2019, LE increased in the general Scottish population and the LE gap widened for people with schizophrenia. For 2000-2002, men and women with schizophrenia lost an excess 9.4 (95% CI 8.5 to 10.3) and 8.2 (7.4 to 9.0) life years, respectively, compared to the general population. In 2017-2019, this excess life years lost increased to 11.8 (10.9 to 12.7) and 11.1 (10.0 to 12.1) for men and women, respectively. There was no evidence of a change over time in the LE gap of 5 to 8 years for people with bipolar disorder or major depression. Changes in LE for natural and unnatural causes of death varied by individual SMI and sex.

**Conclusions:** The LE gap in people with an SMI persisted or widened in Scotland from 2000-2019. These entrenched disparities reflect intersecting inequalities requiring coordinated solutions at multiple levels to improve LE in this and other marginalised and socially excluded groups.

**SUMMARY BOX:** *What is already known on this topic:*

- People with a severe mental illness (SMI) such as schizophrenia, bipolar disorder or major depression have a lower life expectancy (LE) than people without these conditions
- This gap is largely due to natural causes, particularly cardiovascular disease
- LE has increased for the general population but it is unclear whether the gaps in life expectancy for people with SMI have changed over time within the UK

*What this study adds:*

- The LE gap widened in people with schizophrenia and persisted for those with bipolar disorder and major depression in Scotland between 2000 and 2019
- These findings reflect entrenched disparities in the LE of people with SMI
- Multi-level and synergistic solutions are needed to address these disparities

## INTRODUCTION

Since 2010, improvement in life expectancy in the UK general population has stalled.^1–4^ This has been accompanied by widening gaps in life expectancy, such as that observed for people living in the most versus least deprived areas.^2^ ^3^ ^5^ ^6^ It is, however, unclear how such trends have affected the long-recognised yet stubbornly high premature mortality among people with severe mental illness (SMI). Some studies, including from the UK, report a widening in the *relative* mortality gap among those with SMI compared to the general population in recent decades, based on analyses of measures such as standardised mortality ratios.^7–10^ However, from a public health impact perspective, it is important to examine changes in *absolute* measures of excess mortality, such as life expectancy. Recent meta-analyses report average life expectancy disparities of 14.5 years for schizophrenia,^11^ 12.9 years for bipolar disorder,^12^ and 7-19 years for clinically diagnosed depression,^13^ with substantial variation in individual estimates. These variations are likely to be attributable to differences in settings (including health care systems), time periods, and methods of calculating life expectancy. There is limited data on life expectancy of people with SMI in the UK, with most estimates based on data from South London^14–16^ and the only national estimates derived from a Scottish study spanning the period from 1986 to 2010.^17^

Only one UK-based study, from South London, has examined time-trends in life expectancy for people with bipolar disorder and schizophrenia, with the authors reporting an overall small narrowing of life expectancy disparities between 2008-2012 and 2013-2017, with variation by disorder and sex.^14^ Studies outside the UK are conflicting, suggesting widening,^18^ narrowing,^19^ or no change^20–23^ in mortality gaps over time. Many of these studies included non-contemporaneous time periods and most included composite mood disorder groups, with few examining bipolar disorder and major depression separately. Additionally, most existing studies have assumed a fixed age of onset of SMI (often at birth or age 15 years), which does not account for the distribution of age at onset of SMI and potentially biases life expectancy estimates.^24^

In light of evolving methodological approaches to calculating life expectancy^24^ and the limited study of life expectancy time-trends in people with SMI in the UK, we aimed to determine life years lost among people with SMI (defined here as schizophrenia, bipolar disorder or major depression) in Scotland, stratified by sex, from 2000-2019 for all deaths and natural and unnatural deaths. We also assessed whether the life expectancy gap for SMI compared to the general population has changed over time.

## METHODS

We have reported this study in accordance with the Reporting of Studies Conducted using Observational Routinely-Collected Health Data (RECORD) statement.^25^

We obtained permission to conduct this study with pseudonymised, non-consented data from the Public Benefit and Privacy Panel of National Health Service National Services Scotland, reference number 1516-0626.

### Data

The electronic Data Research and Innovation Service (eDRIS) provided us with records from the Scottish Morbidity Record SMR04 dataset, which includes information on admissions to all NHS psychiatric hospitals in Scotland.^26^ We included adults aged at least 18 years old admitted from 1 January 1981 to 31 December 2019. SMR04 includes admission date; age, sex and area-based deprivation (measured by the Carstairs Index^27^ ^28^) at time of admission; and up to six discharge diagnoses. Diagnoses were coded using International Classification of Diseases (ICD)-version 9 up to March 1996 and ICD-10 from April 1996. The eDRIS team linked the SMR04 data via individuals’ unique community health index number to National Records of Scotland (NRS) death records, which include date and cause of death from the death certificate. We sourced life expectancy data for the whole Scottish population from publicly available life tables produced by NRS.^29^

### Severe mental illness

We identified people with a psychiatric hospital diagnosis of schizophrenia (ICD-10 F20 and F25; ICD-9 295.0–295.3 and 295.6–295.9), bipolar disorder (ICD-10 F30-F31; ICD-9 296.0 and 296.2–296.6), or major depression (ICD-10 F32-F33; ICD-9 296.1, 298.0, 300.4 and 311). For people with a record of more than one SMI, we used their most severe condition, with schizophrenia considered the most severe, followed by bipolar disorder and major depression. Our study included people who were alive on 1 January 2000, with individuals entering the cohort on their date of first diagnosis of schizophrenia, bipolar disorder or depression.

### Cause of death

We used ICD-10 codes to categorise cause of death into natural and unnatural causes of death. Unnatural causes of death included suicide, self-harm and injuries of undetermined intent (X60-X84, Y10-Y34, Y87.0 and Y87.2), accidents (V01-X59, Y85-Y86) and all other external causes (Y40-Y84, X85-Y09, Y87.1, Y88-89). All other deaths were classified as natural causes.

### Life expectancy

Previously, estimates of life expectancy for conditions not present at birth assumed a fixed age of onset for all people with the condition, with life expectancy calculated using population mortality rates up to the fixed age, and condition-specific mortality rates beyond the fixed age.^24^ Many studies of life expectancy for people with SMI have used population mortality rates up to 15 years of age and condition-specific mortality rates beyond.^14–16^ ^18^ ^30^ ^31^ However, a limitation of this approach is that it does not account for variation in the age of onset and can hence lead to biased estimates of life expectancy and years of life lost due to the condition.^24^ In the present study we used a recently developed method for calculating the years of life lost due to a condition, termed the Life Years Lost (LYL) method.^24^ ^32^ Unlike earlier methods, this accounts for variation in age of onset of the condition by estimating the LYL for each possible age of onset up to a set maximum age and calculating a weighted average of LYL across the ages of onset. Where the probability of survival beyond the set maximum age is low, the LYL can be interpreted as the average reduction in life expectancy due to the condition. We chose a maximum age of 95 since few people in Scotland survive beyond this age^33^ and to ensure consistency with previous studies.^21^ ^23^ ^34^

### Statistical analysis

For each combination of SMI and sex we summarised age, calendar year and area-based deprivation at the time of first SMI diagnosis as well as the number of deaths, age at death and cause of death.

We calculated LYL separately for each SMI and sex. To evaluate trends over time, we calculated rolling averages of LYL for 18 overlapping 3-year periods from 2000-2002 to 2017-2019, aligning with time periods for official estimates of life expectancy in the Scottish population.^29^ For each period, we created a sub-cohort of people who were alive at the start of the 3-year period and had a first record of SMI diagnosis either before or during the 3-year period. We defined age at start of follow-up as: age on 1 January of the index year for people with an SMI diagnosis record prior to the start of the 3-year period; or age at diagnosis for people with a first-recorded SMI diagnosis during the 3-year period. We identified deaths during the 3-year period, and defined age at the end of follow-up as: age on 31 December of the final year for people who did not die within the 3-year period; and age at the time of death for people who died within the period. For each combination of SMI, sex and period, we calculated LYL between the ages of 18 and 95 years, and decomposed the results into LYL due to natural and unnatural causes.^35^ For each individual year of age from 18 to 94 years, we then estimated the remaining life expectancy overall and by natural or unnatural cause of death and used bootstrapping to estimate confidence intervals. Finally, we estimated overall LYL between the ages of 18 and 95 years by weighting the age-specific estimates of remaining life expectancy according to age at first-recorded SMI diagnosis. For each combination of SMI and sex, we calculated the age at SMI diagnosis distribution based on the entire cohort, rather than the period-specific cohort. This approach ensured that the distribution was based on a large sample, allowing trends in LYL over time to solely reflect changes in life expectancy rather than changes in both life expectancy and the distribution of age at SMI diagnosis.

We calculated excess life years lost for people with SMI compared to the Scottish population, for each combination of SMI, sex, 3-year calendar period, and age between 18 and 95 years, using data from the Scottish life tables. As above, the overall excess LYL was estimated by weighting the age-specific excess LYLs by the age at SMI diagnosis distribution. Hence, the overall excess LYL can be interpreted as the additional LYLs from diagnosis by people with an SMI relative to a population of the same age.

To examine survival patterns by age, we plotted survival curves for each SMI and the Scottish population by sex for the earliest (2000-2002) and latest (2017-2019) periods. We plotted survival curves from age 35 to ensure a sufficient number of people with SMI at risk at each age.

We used R version 4.2.0^36^ to conduct all data preparation, analysis and visualisation, and specifically the R package lillies^24^ to implement the LYL method. The R code is available on GitHub: https://github.com/kjfleetwood/le_smi_scot.

### Patient and public involvement

No patients were involved in the design, conduct or interpretation of this study.

## RESULTS

Our cohort of people with SMI included 117,958 people (fig 1), of whom 28,797 (24.4%) had schizophrenia, 16,657 (14.1%) had bipolar disorder and 72,504 (61.5%) had major depression (table 1). Age at first-recorded SMI diagnosis was lowest amongst people with schizophrenia, with a median of 31 years for men and 37 years for women. For bipolar disorder and depression, the median age of admission was approximately 40 years for both men and women. People with schizophrenia or major depression as primary diagnoses generally lived in more deprived, but there was no association between deprivation and bipolar disorder. Approximately one third of the cohort died during the study period, with median age at death lower for people with schizophrenia (60 years for men, and 71 years for women) than those with bipolar disorder (69 years for men, and 75 years for women) or major depression (70 years for men, and 77 years for women).

**Figure 1:**
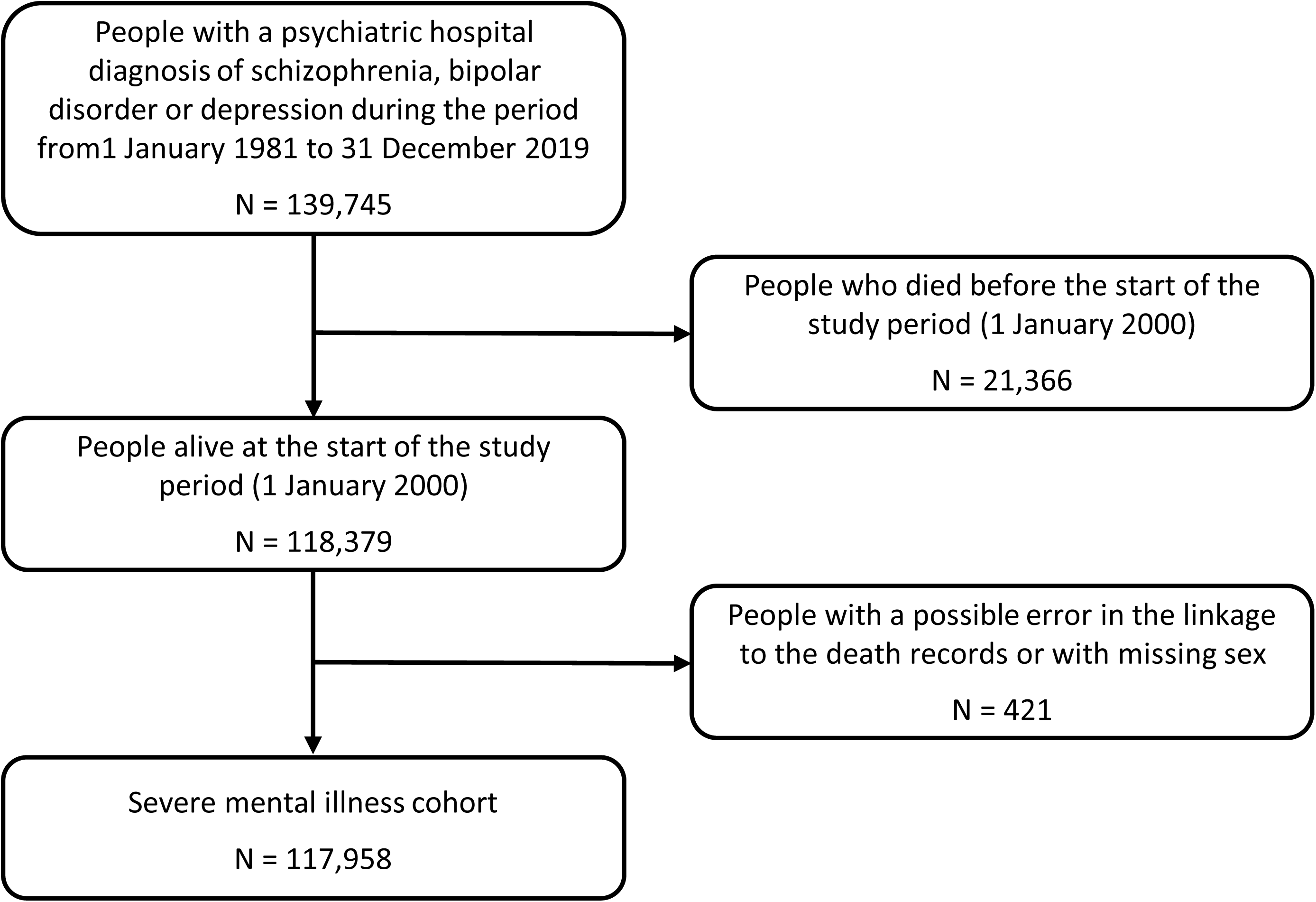
Flow diagram for the severe mental illness cohort

**Table 1:** Baseline characteristics and deaths in the cohort by severe mental illness and sex.

|  | Schizophrenia<br>Male<br>(N=17492) | Schizophrenia<br>Female<br>(N=11305) | Bipolar disorder<br>Male<br>(N=6420) | Bipolar disorder<br>Female<br>(N=10237) | Major<br>Depression<br>Male<br>(N=29379) | Major<br>Depression<br>Female<br>(N=43125) |
| --- | --- | --- | --- | --- | --- | --- |
| **Age at first-recorded SMI diagnosis [Median (Q1, Q3)]** | 31.0<br>(24.0, 41.0) | 37.0<br>(28.0, 51.0) | 40.0<br>(29.0, 53.0) | 41.0<br>(30.0, 54.0) | 42.0<br>(31.0, 56.0) | 43.0<br>(31.0, 59.0) |
| **Year of first-recorded SMI diagnosis\*** |  |  |  |  |  |  |
| 1981-1989 | 4710 (26.9%) | 4274 (37.8%) | 1430 (22.3%) | 3116 (30.4%) | 4795 (16.3%) | 10749 (24.9%) |
| 1990-1999 | 5313 (30.4%) | 3083 (27.3%) | 1770 (27.6%) | 2656 (25.9%) | 8520 (29.0%) | 13008 (30.2%) |
| 2000-2009 | 4512 (25.8%) | 2383 (21.1%) | 1753 (27.3%) | 2409 (23.5%) | 9273 (31.6%) | 11983 (27.8%) |
| 2010-2019 | 2957 (16.9%) | 1565 (13.8%) | 1467 (22.9%) | 2056 (20.1%) | 6791 (23.1%) | 7385 (17.1%) |
| **Carstairs Index quintile at time of SMI diagnosis** |  |  |  |  |  |  |
| 1 (most deprived) | 5026 (28.7%) | 3049 (27.0%) | 1251 (19.5%) | 2078 (20.3%) | 7357 (25.0%) | 10841 (25.1%) |
| 2 | 3662 (20.9%) | 2374 (21.0%) | 1233 (19.2%) | 2006 (19.6%) | 6098 (20.8%) | 9049 (21.0%) |
| 3 | 2988 (17.1%) | 2042 (18.1%) | 1181 (18.4%) | 1930 (18.9%) | 5650 (19.2%) | 8281 (19.2%) |
| 4 | 2502 (14.3%) | 1793 (15.9%) | 1147 (17.9%) | 1861 (18.2%) | 5061 (17.2%) | 7317 (17.0%) |
| 5 (least deprived) | 2050 (11.7%) | 1512 (13.4%) | 1261 (19.6%) | 1959 (19.1%) | 4301 (14.6%) | 6649 (15.4%) |
| Missing | 1264 (7.2%) | 535 (4.7%) | 347 (5.4%) | 403 (3.9%) | 912 (3.1%) | 988 (2.3%) |
| **Deaths [N (%)]** | 5026 (28.7%) | 3991 (35.3%) | 1966 (30.6%) | 3270 (31.9%) | 10341 (35.2%) | 15467 (35.9%) |
| **Age at death [Median (interquartile range)]** | 60.0<br>(47.0, 71.0) | 71.0<br>(60.0, 80.0) | 69.0<br>(57.0, 78.0) | 75.0<br>(65.0, 83.0) | 70.0<br>(55.0, 80.0) | 77.0<br>(64.0, 85.0) |
| **Cause of death [N (% of deaths)]** |  |  |  |  |  |  |
| Natural | 4127 (82.1%) | 3648 (91.4%) | 1691 (86.0%) | 2962 (90.6%) | 8765 (84.8%) | 14126 (91.3%) |
| Unnatural | 899 (17.9%) | 343 (8.6%) | 275 (14.0%) | 308 (9.4%) | 1576 (15.2%) | 1341 (8.7%) |

\*Indicates year of first psychiatric hospital record of SMI

Amongst both men and women, people with schizophrenia lost more life years than people with bipolar disorder or major depression and across all SMIs more life years were lost due to natural rather than unnatural causes (fig 2, supplementary table 1). For schizophrenia, there was no evidence of a change in life expectancy between 2000 and 2019. Compared to a maximum age of 95 years, men with schizophrenia lost an average of 28.9 (95% confidence interval 28.0 to 29.8) years in 2000-2002 and 28.1 (27.2 to 29.0) years in 2017-2019; women lost an average of 22.2 (21.4 to 23.0) years in 2000-2002, and 23.2 (22.1 to 24.3) years in 2017-2019. For men with schizophrenia, LYL due to natural causes decreased from 24.5 (23.6, 25.4) years in 2000-2002 to 21.8 (20.9, 22.7) years in 2017-2019, whilst LYL due to unnatural causes increased from 4.4 (3.5 to 5.2) years to 6.3 (5.3 to 7.3) years. For women with schizophrenia, there was no evidence of a change in LYL due to either natural or unnatural causes.

**Figure 2:**
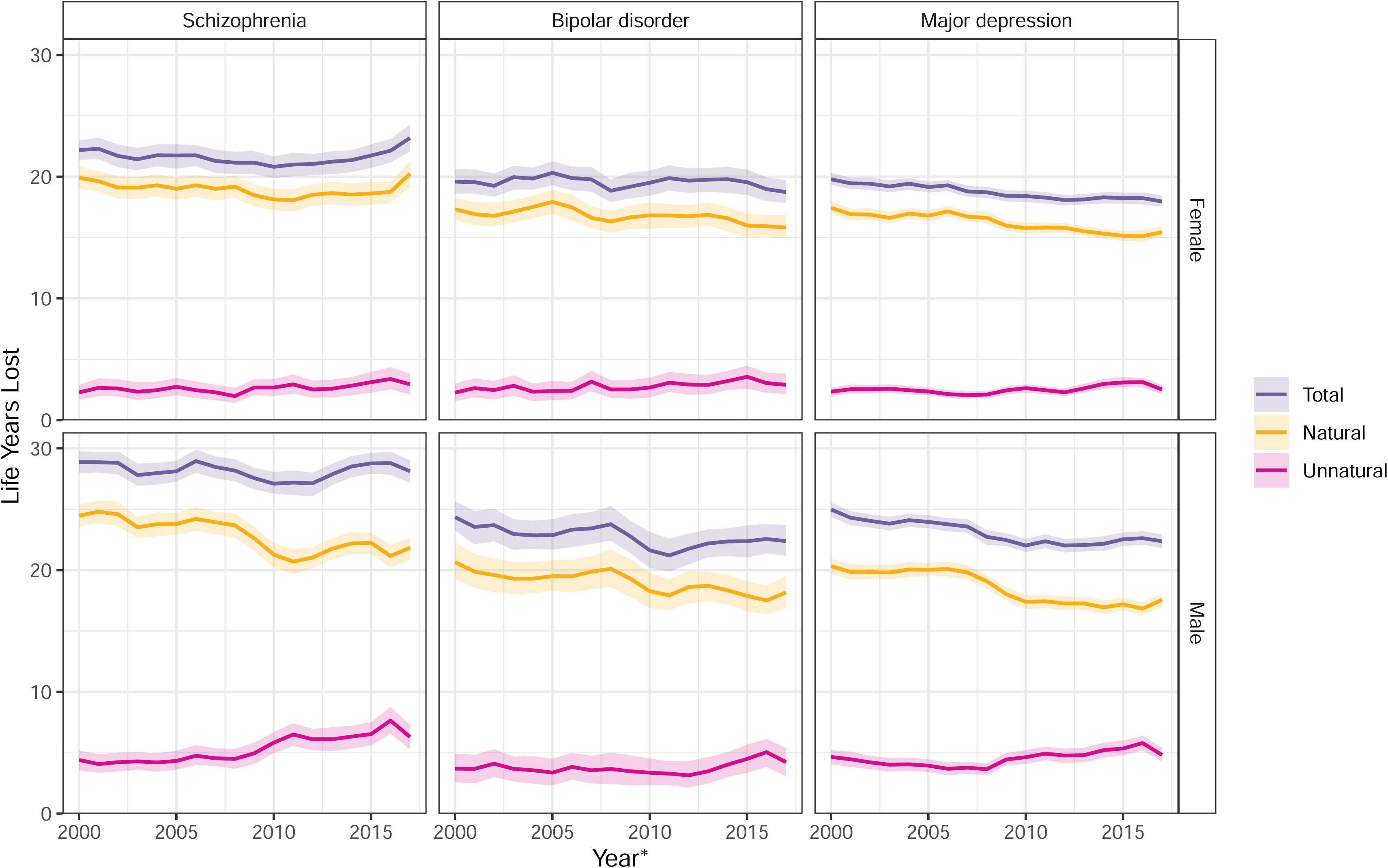
Life years lost between 18 and 95 years of age for all deaths and natural and unnatural deaths among people in Scotland with each severe mental illness, stratified by sex, (rolling three-year averages between 2000 and 2019) at the start of three-year period

Life expectancy improved among both men and women with major depression between 2000 and 2019. Compared to a maximum age of 95 years, men with major depression lost an average of 25.0 (24.4 to 25.5) years in 2000-2002 and 22.4 (21.8 to 22.9) years in 2017-2019, whilst women lost an average of 19.8 (19.4 to 20.3) years in 2000-2002, and 18.0 (17.5 to 18.5) years in 2017-2019. These improvements stemmed from reductions in LYL due to natural causes, with no observed change in LYL due to unnatural causes.

The pattern for people with bipolar disorder was less clear given the smaller numbers in this group. For both men and women, life expectancy may have improved slightly between 2000 and 2019, but the confidence intervals overlapped. Compared to a maximum age of 95 years, men with bipolar disorder lost an average of 24.4 (23.3 to 25.7) years in 2000-2002 and 22.4 (21.2 to 23.7) years in 2017-2019, whilst women lost an average of 19.6 (18.6 to 20.6) years in 2000-2002, and 18.7 (17.9 to 19.7) years in 2017-2019. For both men and women any improvement in life expectancy appears to be driven by a reduction in LYL due to natural rather than unnatural causes of death.

Overall, life expectancy in the Scottish population increased between 2000 and 2019.^37^ For men, life expectancy increased from 73.3 years in 2000-2002 to 77.1 years in 2017-2019, whilst for women it increased from 78.8 in 2000-2002 to 81.1 years in 2017-2019. Compared with the Scottish population, people with each SMI had lower life expectancies (fig 3, supplementary table 2). The gap in life expectancy for people with schizophrenia increased between 2000 and 2019. In 2000-2002, men with schizophrenia lost 9.4 (95% confidence interval 8.5 to 10.3) excess life years from diagnosis compared with men of the same age from the Scottish population. This gap had increased by 2017-2019 with men with schizophrenia losing 11.8 (10.9 to 12.7) excess life years. Similarly, women with schizophrenia lost 8.2 (7.4 to 9.0) excess life years in 2000-2002 and 11.1 (10.0 to 12.1) excess life years in 2017-2019.

**Figure 3:**
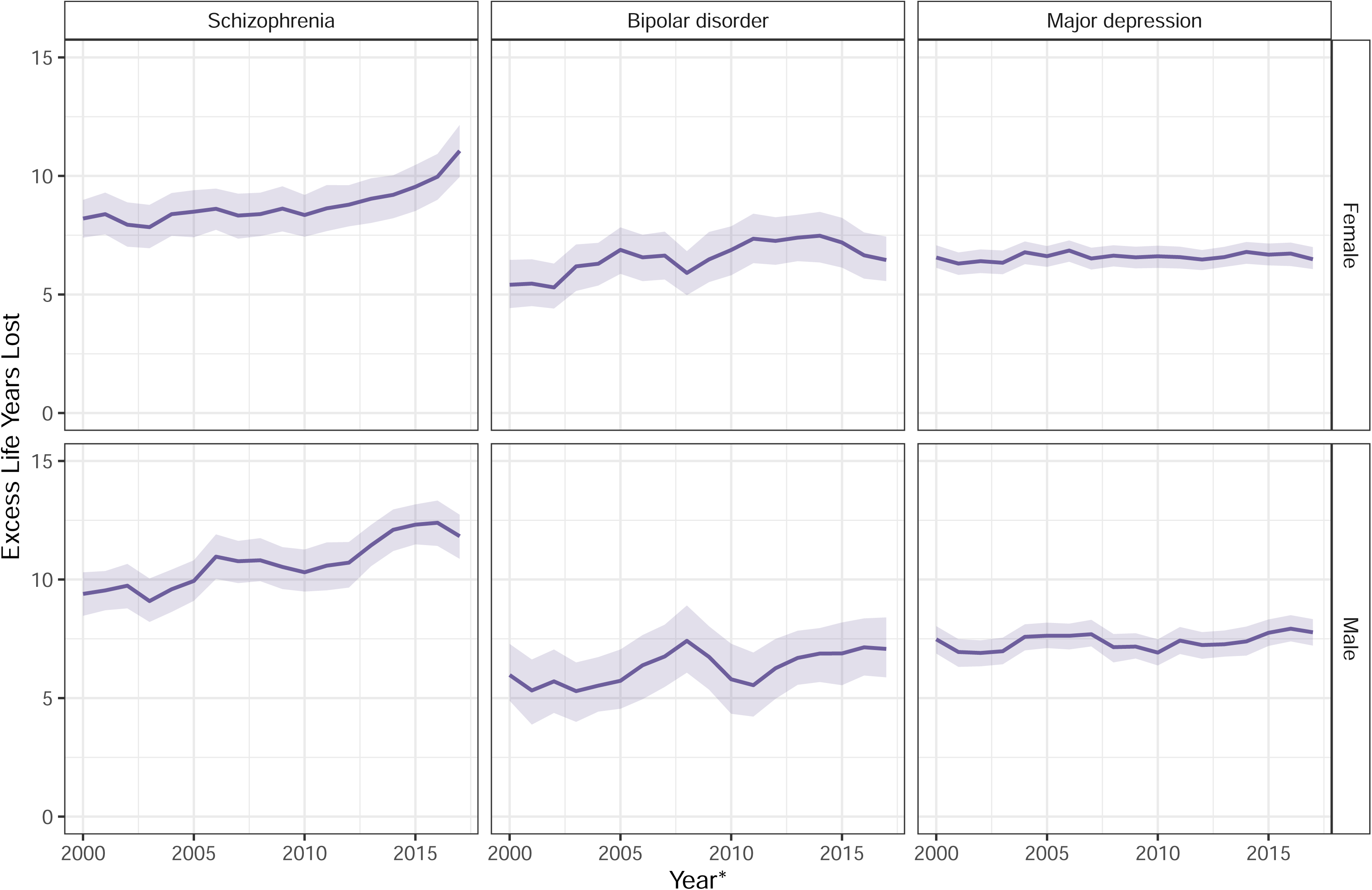
Excess life years lost in people with a severe mental illness in comparison to the Scottish population, stratified by sex (rolling three-year averages between 2000 and 2019) * at the start of three-year period

For bipolar disorder and major depression, there was no evidence of a change in the life expectancy gap relative to the Scottish population between 2000 to 2019. Men with bipolar disorder lost 6.0 (4.9 to 7.3) excess life years in 2000-2002 and 7.1 (5.9 to 8.4) excess life years in 2017-2019. Women with bipolar disorder lost 5.4 (4.4 to 6.5) excess life years in 2000-2002 and 6.5 (5.6 to 7.4) excess life years in 2017-2019. The gap was similar for people with major depression, with men losing 7.5 (6.9 to 8.0) excess life years in 2000-2002 and 7.8 (7.2 to 8.3) excess life years in 2017-2019. Women with major depression lost 6.6 (6.1 to 7.1) excess life years in 2000-2002 and 6.5 (6.1 to 7.0) excess life years in 2017-2019.

In both the earliest (2000-2002) and most recent (2017-2019) periods, differences in survival between the Scottish population and people with SMI are evident across the life span from 35 to 95 years (fig 4).

**Figure 4:**
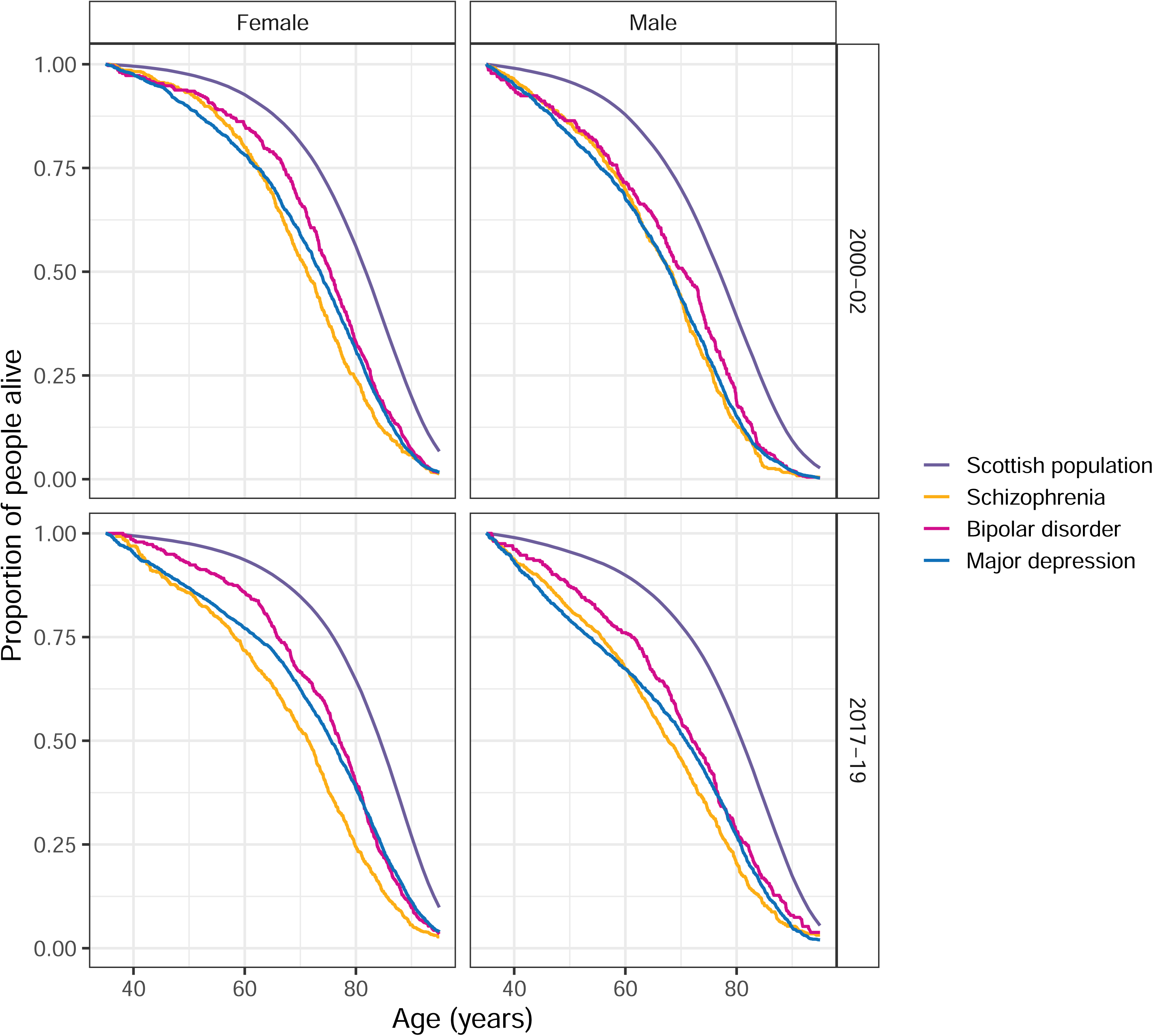
Survival curves for the Scottish population and those with each of schizophrenia, bipolar disorder and major depression, stratified by sex, in 2000-2002 and 2017-2019

## DISCUSSION

Our findings reveal persistent large disparities in the life expectancy of those with SMI compared to the general population that are evident across the entire lifespan. In Scotland, between 2000 and 2019, life expectancy in the general population increased. Although life expectancy similarly improved among those with a psychiatric hospital admission record for major depression (driven by reductions in LYL due to natural causes) the life expectancy gap did not narrow. In contrast, the gap for people with schizophrenia appears to have widened. Among men with schizophrenia, LYL due to natural causes decreased, whilst LYL due to unnatural causes increased, with no changes observed among women. The pattern is less clear for bipolar disorder (due to small numbers in this sub-population) and whilst life expectancy may have improved slightly, there is no evidence that the gap is narrowing.

### Comparison with other studies

Our estimates of the life expectancy gap for each of schizophrenia, bipolar disorder and major depression are lower than reported in some previous studies, including UK studies.^11^ ^12^ ^14^ ^17^ This may reflect differences in study setting, period and methodology, including our adoption of the new LYL method, which takes account of the variation in age of onset of psychiatric illness, providing more conservative life expectancy estimates.^24^ Our estimates generally align with other recent studies from Denmark and Hong Kong that also used the LYL method.^20–23^ ^34^

In contrast to our study, previous contemporaneous studies from Denmark and Hong Kong employing the LYL method found no change in the gap over time for people with schizophrenia compared to the general population.^21–23^ Where mood disorders were analysed, studies did not disaggregate bipolar disorder and major depression,^22^ but estimates of life expectancy and lack of change in the gap over time aligned with our findings. Interestingly, where changes in groups of causes of death were examined in previous Danish studies, authors found that reductions in unnatural causes of death were offset by increasing deaths from natural causes.^20–22^ This contrasts with our findings, where, although patterns varied by sex and mental disorder, we generally found a decrease in LYL from natural causes of death and no change or an increase in LYL from unnatural causes of deaths. This may reflect the under-funding of mental health services in the UK during this time period.^38^ The only other contemporary UK-based study, based in south London, found a narrowing of the life expectancy gap from 2008-2012 to 2013-2017 for all SMI disorders combined.^14^ However, in this study confidence intervals of estimates for the two periods overlapped and findings differed by mental disorders, with an improvement in life expectancy for men (but not women) with schizophrenia, a possible worsening of the gap for men with bipolar disorder and no change among women with bipolar disorder. This study included a single highly urbanised area, which limits its generalisability to the rest of England and the wider UK. The authors also used UK-wide life expectancy estimates rather than comparing SMI life expectancy to that of the defined geographical area of the study population. Our findings differ from those of two studies based on earlier decades of data. Lawrence et al, reported widening life expectancy gap for mental disorders compared to the general population in Australia from 1985-2005,^18^ whereas Wahlbeck et al. reported a very slight narrowing of the life expectancy gap for some, but not all Nordic countries from 1987-2006.^19^ These apparent inconsistencies may reflect the differing time periods of interest and settings, including different health systems, as well as differing methodology.

Mortality rate ratios in people with schizophrenia and bipolar disorder relative to that of the general population have been reported to be widening between 2000 and 2014 in the UK.^7^ Relative measures are sensitive to decreases in the reference population mortality rates over time, resulting in larger relative differences between groups; but are essentially consistent with the changes in absolute differences in life expectancy over time that we have reported.

### Strengths and limitations of this study

A key strength of our study is the use of recent national level data spanning two decades, which enabled assessment of change in life expectancy over a substantial period of time with stratification by sex. Given the use of whole population data during a long study period we were able to analyse life expectancy by individual SMI, and in particular distinguish between bipolar disorder and major depression, which have often been combined in a composite group of all mood disorders in previous studies, potentially masking differing patterns by disorder. Finally, we adopted the recently introduced LYL method that accounts for the age of diagnosis distribution and allows decomposition of life years lost by cause of death. Given there is sometimes a delay between onset of psychiatric disorder and diagnosis, the true gap in life expectancy for those with SMI is likely to lie between estimates generated by this more conservative approach and earlier, less conservative, methods. Our study has some limitations. In the absence of availability of national primary care data for research purposes or a mental health registry, we were only able to ascertain SMI through hospital admission data. We are therefore likely to have included people with SMI at the more severe end of the spectrum or with particularly challenging/complex health and social care needs. Therefore, our findings may not be generalisable to all people with SMI. Moreover, we used first hospital admission date as a proxy of diagnosis date and so estimates of life expectancy from disease onset may be slightly under-estimated. It is also possible that the widening gap in LE for people with schizophrenia reflects changes in practice related to psychiatric hospital admission in Scotland, with number of psychiatric hospital admissions having steadily decreased from around 30,000 in 2000 to around 20,000 from 2010 onwards.^39^ It is therefore possible that people admitted in later years have more severe illness than those admitted in earlier years, which may contribute to the widening gap.

Although we included nationwide data, the number of people included with bipolar disorder was smaller than in the other groups and LYL and excess LYL estimates were therefore more uncertain. Whilst our study makes a valuable contribution to understanding the contemporary disparity in life expectancy of people with SMI, a number of questions were beyond the scope of our study and warrant investigation. These include: further examination of the role of deprivation, with previous studies indicating an effect of SMI on excess mortality that is over and above that of deprivation;^16^ closer investigation of changes in cause-specific mortality over time, particularly alcohol-related deaths;^13^ the role of changes in mental health services (with fewer inpatient beds available); and the impact of the COVID-19 pandemic and its aftermath on life expectancy among those with SMI, given reports of higher COVID-19 mortality in people with SMI.^40^

### Implications

Excess deaths due to natural causes account for much of the observed premature mortality in people with SMI in both our and a Danish study^20^ reflecting the poorer physical health of people with SMI, in whom there is a high burden of multimorbidity.^41^ The cause of this premature mortality is complex, multifactorial and multi-level,^42^ ^43^ with key risk factors operating at the individual through to societal level and encompassing: modifiable lifestyle behaviours such as smoking, overweight/obesity and physical inactivity; multimorbidity and substance misuse; psychological factors; socioeconomic disadvantage; health care access, including year-on-year under-funding of mental health services, and diagnostic overshadowing; iatrogenic harms of some psychotropic medication; structural inequalities; and stigma and marginalisation.^43^ Addressing these complex causes require multi-level and synergistic solutions.^42^ ^43^ These have recently been structured around three key principles: integration of mental and physical health care; prioritisation of prevention of physical disease whilst strengthening treatment of mental illness; and optimisation of intervention synergies across social-ecological levels.^43^ Current health system structures generally create disconnected care that particularly disadvantages patients with complex needs, such as those with SMI. In particular there is a need for better integration across primary, secondary and community health care settings and between generalist and specialist providers, along with co-ordinated support from social care and allied health professionals^42^ ^44^ In addition to regular physical health monitoring in those with SMI, a greater focus on primary prevention strategies is required to tackle key modifiable risk behaviours in this group.^45^

### Conclusion

Our national study highlights the persistent and potentially widening gap in life expectancy experienced by people with SMI. This entrenched disparity reflects ingrained inequalities at multiple levels that affect the health and lifespan of this marginalised sub-group of the population. Future success in narrowing this shameful gap relies on government commitment to introducing equitable social, economic and health policies, significant healthcare re-structure and improved resourcing, and investment in tailored, well-implemented and appropriately evaluated interventions and strategies for primary and secondary prevention of SMI and associated co-morbidities.

### Ethical approval

Permission to conduct this study with pseudonymised, non-consented data was obtained from the Public Benefit and Privacy Panel of National Health Service National Services Scotland, reference number 1516-0626.

## Supporting information

RECORD Checklist

## Data Availability

Researchers can request access to the Scottish records used in this analysis by contacting the electronic Data Research and Innovation Service (eDRIS). Details of the available datasets and the application process are available from https://publichealthscotland.scot/services/data-research-and-innovation-services/electronic-data-research-and-innovation-service-edris. R scripts for the analyses presented in the paper are available at: https://github.com/kjfleetwood/le_smi_scot.

## Acknowledgements

The authors would like to acknowledge the support of the eDRIS Team (Public Health Scotland) for their involvement in obtaining approvals, provisioning and linking data and the use of the secure analytical platform within the National Safe Haven.

## Footnotes

### Contributors

CJ developed the idea for the study and obtained funding. RA and KF conducted the literature review. KF prepared the data and conducted the statistical analyses. All authors reviewed and interpreted the results. CJ and KF wrote the initial draft of the manuscript. All authors revised the manuscript and approved the final version. CJ acts at guarantor for the paper. The corresponding author attests that all listed authors meet authorship criteria and that no others meeting the criteria have been omitted.

## Transparency statement

The guarantor (CJ) affirms that the manuscript is an honest, accurate, and transparent account of the study being reported; that no important aspects of the study have been omitted; and that any discrepancies from the study as originally planned have been explained.

## Dissemination to participants and related patient and public communities

Findings from the current work will be disseminated through open access publication at *The BMJ*, the authors’ institutional and personal social media accounts as appropriate, and potential press releases.

## Copyright/license for publication

The Corresponding Author has the right to grant on behalf of all authors and does grant on behalf of all authors, an exclusive licence on a worldwide basis to the BMJ Publishing Group Ltd to permit this article (if accepted) to be published in BMJ editions and any other BMJPGL products and sublicences such use and exploit all subsidiary rights, as set out in our licence.

