## Supplement for "Time-trends in life expectancy of people with severe mental illness in Scotland, 2000-2019: a population-based study"

### Supplementary Materials

**Table of Contents**

Supplementary Table 1: Life years lost between 18 and 95 years of age for all deaths and natural and unnatural deaths among people in Scotland with each severe mental illness, stratified by sex (rolling three-year averages between 2000 and 2019) ........................................................................................ 2

Supplementary Table 2: Excess life years lost in people with a severe mental illness in comparison to the Scottish population, stratified by sex (rolling three-year averages between 2000 and 2019)........ 5

**Supplementary Table 1: Life years lost between 18 and 95 years of age for all deaths and natural and unnatural deaths among people in Scotland with each severe mental illness, stratified by sex (rolling three-year averages between 2000 and 2019)**

| | | | SMI | | |
|---|---|---|---|---|---|
| Cause of death | Sex | Period | Schizophrenia | Bipolar disorder | Major depression |
| All | Male | 2000-02 | 28.9 (28.0, 29.8) | 24.4 (23.3, 25.7) | 25.0 (24.4, 25.5) |
| | | 2001-03 | 28.9 (28.0, 29.7) | 23.5 (22.1, 24.9) | 24.3 (23.7, 24.8) |
| | | 2002-04 | 28.8 (27.9, 29.7) | 23.7 (22.4, 25.0) | 24.0 (23.5, 24.6) |
| | | 2003-05 | 27.8 (26.9, 28.8) | 23.0 (21.7, 24.2) | 23.8 (23.3, 24.4) |
| | | 2004-06 | 28.0 (27.0, 28.8) | 22.9 (21.8, 24.1) | 24.1 (23.5, 24.6) |
| | | 2005-07 | 28.1 (27.3, 29.0) | 22.9 (21.7, 24.2) | 24.0 (23.4, 24.5) |
| | | 2006-08 | 29.0 (28.0, 29.9) | 23.3 (21.9, 24.6) | 23.8 (23.2, 24.3) |
| | | 2007-09 | 28.5 (27.6, 29.3) | 23.4 (22.1, 24.8) | 23.6 (23.1, 24.2) |
| | | 2008-10 | 28.2 (27.3, 29.1) | 23.8 (22.4, 25.3) | 22.7 (22.1, 23.3) |
| | | 2009-11 | 27.6 (26.6, 28.4) | 22.8 (21.4, 24.1) | 22.5 (22.0, 23.0) |
| | | 2010-12 | 27.1 (26.3, 28.1) | 21.6 (20.2, 23.1) | 22.0 (21.5, 22.6) |
| | | 2011-13 | 27.2 (26.2, 28.2) | 21.2 (19.9, 22.6) | 22.4 (21.8, 22.9) |
| | | 2012-14 | 27.1 (26.1, 28.0) | 21.8 (20.5, 23.0) | 22.0 (21.4, 22.6) |
| | | 2013-15 | 27.9 (27.0, 28.7) | 22.2 (21.1, 23.3) | 22.1 (21.5, 22.6) |
| | | 2014-16 | 28.5 (27.6, 29.4) | 22.4 (21.2, 23.4) | 22.2 (21.6, 22.8) |
| | | 2015-17 | 28.8 (27.9, 29.6) | 22.4 (21.0, 23.7) | 22.5 (22.0, 23.1) |
| | | 2016-18 | 28.8 (27.8, 29.7) | 22.6 (21.4, 23.8) | 22.6 (22.1, 23.2) |
| | | 2017-19 | 28.1 (27.2, 29.0) | 22.4 (21.2, 23.7) | 22.4 (21.8, 22.9) |
| | Female | 2000-02 | 22.2 (21.4, 23.0) | 19.6 (18.6, 20.6) | 19.8 (19.4, 20.3) |
| | | 2001-03 | 22.3 (21.4, 23.2) | 19.6 (18.6, 20.6) | 19.5 (19.0, 19.9) |
| | | 2002-04 | 21.7 (20.8, 22.6) | 19.2 (18.4, 20.3) | 19.4 (18.9, 19.9) |
| | | 2003-05 | 21.4 (20.5, 22.4) | 20.0 (18.9, 20.9) | 19.2 (18.7, 19.7) |
| | | 2004-06 | 21.8 (20.8, 22.6) | 19.8 (18.9, 20.7) | 19.4 (18.9, 19.9) |
| | | 2005-07 | 21.7 (20.7, 22.6) | 20.3 (19.3, 21.3) | 19.2 (18.7, 19.6) |
| | | 2006-08 | 21.8 (20.9, 22.6) | 19.9 (18.9, 20.8) | 19.3 (18.8, 19.7) |
| | | 2007-09 | 21.3 (20.3, 22.2) | 19.8 (18.8, 20.8) | 18.8 (18.3, 19.2) |
| | | 2008-10 | 21.2 (20.2, 22.1) | 18.8 (17.9, 19.8) | 18.7 (18.3, 19.2) |
| | | 2009-11 | 21.2 (20.2, 22.1) | 19.2 (18.2, 20.3) | 18.4 (18.0, 18.9) |
| | | 2010-12 | 20.8 (19.9, 21.7) | 19.5 (18.4, 20.5) | 18.4 (17.9, 18.9) |
| | | 2011-13 | 21.0 (20.0, 22.0) | 19.9 (18.9, 20.9) | 18.3 (17.8, 18.7) |
| | | 2012-14 | 21.0 (20.1, 21.9) | 19.7 (18.7, 20.7) | 18.1 (17.6, 18.5) |
| | | 2013-15 | 21.2 (20.2, 22.1) | 19.8 (18.8, 20.7) | 18.1 (17.7, 18.6) |
| | | 2014-16 | 21.4 (20.4, 22.2) | 19.8 (18.7, 20.8) | 18.3 (17.8, 18.7) |
| | | 2015-17 | 21.7 (20.7, 22.7) | 19.6 (18.5, 20.6) | 18.2 (17.8, 18.7) |
| | | 2016-18 | 22.1 (21.2, 23.1) | 19.0 (18.0, 19.9) | 18.2 (17.7, 18.7) |
| | | 2017-19 | 23.2 (22.1, 24.3) | 18.7 (17.9, 19.7) | 18.0 (17.5, 18.5) |

| | | | SMI | | |
|---|---|---|---|---|---|
| Cause of death | Sex | Period | Schizophrenia | Bipolar disorder | Major depression |
| Natural | Male | 2000-02 | 24.5 (23.6, 25.4) | 20.7 (19.3, 22.3) | 20.3 (19.7, 20.9) |
| | | 2001-03 | 24.8 (23.9, 25.7) | 19.9 (18.5, 21.3) | 19.8 (19.2, 20.5) |
| | | 2002-04 | 24.6 (23.6, 25.6) | 19.6 (18.2, 20.9) | 19.8 (19.3, 20.4) |
| | | 2003-05 | 23.5 (22.6, 24.4) | 19.3 (18.0, 20.7) | 19.8 (19.3, 20.4) |
| | | 2004-06 | 23.8 (22.8, 24.8) | 19.3 (18.1, 20.7) | 20.1 (19.4, 20.7) |
| | | 2005-07 | 23.8 (22.9, 24.7) | 19.5 (18.3, 20.8) | 20.0 (19.5, 20.6) |
| | | 2006-08 | 24.2 (23.2, 25.2) | 19.5 (18.2, 20.8) | 20.1 (19.5, 20.7) |
| | | 2007-09 | 23.9 (23.0, 24.8) | 19.9 (18.4, 21.2) | 19.8 (19.2, 20.4) |
| | | 2008-10 | 23.7 (22.7, 24.7) | 20.1 (18.6, 21.7) | 19.1 (18.5, 19.6) |
| | | 2009-11 | 22.6 (21.6, 23.5) | 19.3 (17.8, 20.9) | 18.0 (17.5, 18.6) |
| | | 2010-12 | 21.3 (20.3, 22.2) | 18.3 (16.9, 19.8) | 17.4 (16.8, 17.9) |
| | | 2011-13 | 20.7 (19.7, 21.7) | 17.9 (16.7, 19.2) | 17.4 (16.9, 18.0) |
| | | 2012-14 | 21.0 (20.1, 21.9) | 18.6 (17.3, 19.9) | 17.3 (16.7, 17.8) |
| | | 2013-15 | 21.8 (20.9, 22.6) | 18.7 (17.5, 19.9) | 17.3 (16.7, 17.8) |
| | | 2014-16 | 22.2 (21.3, 23.1) | 18.3 (17.2, 19.6) | 16.9 (16.4, 17.5) |
| | | 2015-17 | 22.2 (21.3, 23.0) | 17.9 (16.6, 19.1) | 17.2 (16.7, 17.7) |
| | | 2016-18 | 21.2 (20.2, 22.1) | 17.5 (16.3, 18.7) | 16.8 (16.3, 17.4) |
| | | 2017-19 | 21.8 (20.9, 22.7) | 18.2 (16.9, 19.5) | 17.6 (17.0, 18.1) |
| | Female | 2000-02 | 19.9 (19.0, 20.8) | 17.3 (16.4, 18.2) | 17.4 (17.0, 17.9) |
| | | 2001-03 | 19.6 (18.8, 20.5) | 16.9 (16.0, 17.9) | 16.9 (16.4, 17.4) |
| | | 2002-04 | 19.1 (18.2, 20.0) | 16.8 (15.9, 17.9) | 16.9 (16.4, 17.4) |
| | | 2003-05 | 19.1 (18.2, 19.9) | 17.1 (16.2, 18.1) | 16.6 (16.1, 17.1) |
| | | 2004-06 | 19.3 (18.4, 20.2) | 17.5 (16.6, 18.4) | 17.0 (16.5, 17.4) |
| | | 2005-07 | 19.0 (18.1, 19.9) | 17.9 (16.9, 18.9) | 16.8 (16.4, 17.2) |
| | | 2006-08 | 19.3 (18.3, 20.1) | 17.5 (16.4, 18.5) | 17.1 (16.7, 17.6) |
| | | 2007-09 | 19.0 (18.0, 19.9) | 16.6 (15.7, 17.5) | 16.7 (16.3, 17.2) |
| | | 2008-10 | 19.2 (18.3, 20.1) | 16.3 (15.4, 17.2) | 16.6 (16.2, 17.1) |
| | | 2009-11 | 18.5 (17.6, 19.3) | 16.7 (15.7, 17.6) | 16.0 (15.5, 16.4) |
| | | 2010-12 | 18.1 (17.2, 19.0) | 16.8 (15.7, 17.9) | 15.8 (15.3, 16.2) |
| | | 2011-13 | 18.1 (17.1, 18.9) | 16.8 (15.8, 17.7) | 15.8 (15.4, 16.2) |
| | | 2012-14 | 18.5 (17.6, 19.4) | 16.8 (15.8, 17.7) | 15.8 (15.4, 16.2) |
| | | 2013-15 | 18.6 (17.7, 19.6) | 16.9 (16.0, 17.9) | 15.5 (15.1, 15.9) |
| | | 2014-16 | 18.5 (17.6, 19.4) | 16.6 (15.5, 17.5) | 15.3 (14.9, 15.7) |
| | | 2015-17 | 18.6 (17.7, 19.6) | 16.0 (15.0, 17.0) | 15.1 (14.7, 15.6) |
| | | 2016-18 | 18.8 (17.9, 19.6) | 15.9 (15.0, 16.8) | 15.1 (14.7, 15.6) |
| | | 2017-19 | 20.2 (19.2, 21.2) | 15.8 (15.0, 16.8) | 15.4 (15.0, 15.9) |

| | | | SMI | | |
|---|---|---|---|---|---|
| Cause of death | Sex | Period | Schizophrenia | Bipolar disorder | Major depression |
| Unnatural | Male | 2000-02 | 4.4 (3.5, 5.2) | 3.7 (2.6, 4.9) | 4.7 (4.0, 5.2) |
| | | 2001-03 | 4.1 (3.3, 4.9) | 3.7 (2.5, 4.8) | 4.5 (3.8, 5.1) |
| | | 2002-04 | 4.2 (3.5, 5.0) | 4.1 (3.0, 5.3) | 4.2 (3.7, 4.7) |
| | | 2003-05 | 4.3 (3.6, 5.0) | 3.7 (2.6, 4.7) | 4.0 (3.5, 4.6) |
| | | 2004-06 | 4.2 (3.4, 4.9) | 3.6 (2.5, 4.7) | 4.0 (3.5, 4.6) |
| | | 2005-07 | 4.3 (3.6, 5.2) | 3.4 (2.3, 4.5) | 3.9 (3.4, 4.4) |
| | | 2006-08 | 4.8 (4.0, 5.6) | 3.8 (2.8, 5.0) | 3.7 (3.1, 4.2) |
| | | 2007-09 | 4.5 (3.9, 5.4) | 3.6 (2.5, 4.7) | 3.8 (3.3, 4.3) |
| | | 2008-10 | 4.5 (3.7, 5.3) | 3.7 (2.4, 5.0) | 3.6 (3.1, 4.1) |
| | | 2009-11 | 5.0 (4.1, 5.8) | 3.5 (2.3, 4.8) | 4.4 (4.0, 5.0) |
| | | 2010-12 | 5.8 (5.0, 6.7) | 3.4 (2.3, 4.5) | 4.6 (4.1, 5.2) |
| | | 2011-13 | 6.5 (5.5, 7.4) | 3.3 (2.3, 4.3) | 4.9 (4.4, 5.5) |
| | | 2012-14 | 6.1 (5.2, 6.9) | 3.1 (2.1, 4.3) | 4.8 (4.2, 5.3) |
| | | 2013-15 | 6.1 (5.1, 7.1) | 3.5 (2.5, 4.6) | 4.8 (4.2, 5.4) |
| | | 2014-16 | 6.3 (5.3, 7.2) | 4.0 (3.0, 5.0) | 5.2 (4.6, 5.8) |
| | | 2015-17 | 6.5 (5.6, 7.5) | 4.5 (3.3, 5.7) | 5.3 (4.8, 6.0) |
| | | 2016-18 | 7.6 (6.6, 8.7) | 5.0 (3.8, 6.1) | 5.8 (5.2, 6.4) |
| | | 2017-19 | 6.3 (5.3, 7.3) | 4.2 (3.1, 5.4) | 4.8 (4.3, 5.4) |
| | Female | 2000-02 | 2.3 (1.7, 2.9) | 2.3 (1.6, 3.0) | 2.3 (2.0, 2.7) |
| | | 2001-03 | 2.7 (2.0, 3.4) | 2.6 (1.9, 3.4) | 2.5 (2.2, 2.9) |
| | | 2002-04 | 2.6 (2.0, 3.4) | 2.5 (1.7, 3.2) | 2.5 (2.2, 2.9) |
| | | 2003-05 | 2.3 (1.7, 3.1) | 2.8 (2.0, 3.7) | 2.6 (2.2, 2.9) |
| | | 2004-06 | 2.5 (1.8, 3.2) | 2.3 (1.6, 3.0) | 2.5 (2.1, 2.8) |
| | | 2005-07 | 2.7 (2.1, 3.5) | 2.4 (1.6, 3.2) | 2.4 (2.1, 2.7) |
| | | 2006-08 | 2.5 (1.8, 3.2) | 2.4 (1.7, 3.1) | 2.1 (1.8, 2.5) |
| | | 2007-09 | 2.3 (1.6, 2.9) | 3.2 (2.4, 4.0) | 2.1 (1.8, 2.4) |
| | | 2008-10 | 2.0 (1.4, 2.7) | 2.5 (1.8, 3.3) | 2.1 (1.8, 2.4) |
| | | 2009-11 | 2.7 (2.1, 3.3) | 2.5 (1.8, 3.3) | 2.4 (2.1, 2.8) |
| | | 2010-12 | 2.7 (2.0, 3.4) | 2.7 (1.9, 3.5) | 2.6 (2.2, 2.9) |
| | | 2011-13 | 2.9 (2.2, 3.8) | 3.1 (2.3, 4.0) | 2.5 (2.1, 2.8) |
| | | 2012-14 | 2.5 (1.9, 3.2) | 2.9 (2.1, 3.8) | 2.3 (2.0, 2.6) |
| | | 2013-15 | 2.6 (1.9, 3.3) | 2.9 (2.1, 3.7) | 2.6 (2.3, 3.0) |
| | | 2014-16 | 2.8 (2.0, 3.5) | 3.2 (2.5, 4.0) | 3.0 (2.6, 3.4) |
| | | 2015-17 | 3.1 (2.2, 4.0) | 3.6 (2.6, 4.5) | 3.1 (2.7, 3.5) |
| | | 2016-18 | 3.4 (2.5, 4.3) | 3.0 (2.3, 3.9) | 3.1 (2.7, 3.5) |
| | | 2017-19 | 2.9 (2.1, 3.8) | 2.9 (2.1, 3.8) | 2.5 (2.1, 2.9) |

**Supplementary Table 2: Excess life years lost in people with a severe mental illness in comparison to the Scottish population, stratified by sex (rolling three-year averages between 2000 and 2019)**

| | | SMI | | |
|---|---|---|---|---|
| Sex | Period | Schizophrenia | Bipolar disorder | Depression |
| Male | 2000-02 | 9.4 (8.5, 10.3) | 6.0 (4.9, 7.3) | 7.5 (6.9, 8.0) |
| | 2001-03 | 9.5 (8.7, 10.4) | 5.3 (3.9, 6.6) | 6.9 (6.3, 7.5) |
| | 2002-04 | 9.7 (8.8, 10.7) | 5.7 (4.4, 7.0) | 6.9 (6.3, 7.4) |
| | 2003-05 | 9.1 (8.2, 10.0) | 5.3 (4.0, 6.5) | 7.0 (6.4, 7.5) |
| | 2004-06 | 9.6 (8.6, 10.4) | 5.5 (4.4, 6.7) | 7.6 (7.0, 8.1) |
| | 2005-07 | 9.9 (9.1, 10.8) | 5.7 (4.6, 7.0) | 7.6 (7.1, 8.2) |
| | 2006-08 | 11.0 (10.0, 11.9) | 6.4 (5.0, 7.7) | 7.6 (7.1, 8.1) |
| | 2007-09 | 10.8 (9.9, 11.6) | 6.8 (5.5, 8.1) | 7.7 (7.2, 8.3) |
| | 2008-10 | 10.8 (9.9, 11.7) | 7.4 (6.1, 8.9) | 7.1 (6.5, 7.7) |
| | 2009-11 | 10.5 (9.6, 11.4) | 6.7 (5.4, 8.0) | 7.2 (6.7, 7.7) |
| | 2010-12 | 10.3 (9.5, 11.3) | 5.8 (4.3, 7.3) | 6.9 (6.4, 7.5) |
| | 2011-13 | 10.6 (9.5, 11.6) | 5.5 (4.2, 6.9) | 7.4 (6.9, 8.0) |
| | 2012-14 | 10.7 (9.7, 11.6) | 6.3 (5.0, 7.5) | 7.2 (6.7, 7.8) |
| | 2013-15 | 11.4 (10.6, 12.3) | 6.7 (5.6, 7.8) | 7.3 (6.7, 7.8) |
| | 2014-16 | 12.1 (11.2, 13.0) | 6.9 (5.7, 7.9) | 7.4 (6.8, 8.0) |
| | 2015-17 | 12.3 (11.5, 13.2) | 6.9 (5.5, 8.2) | 7.8 (7.2, 8.3) |
| | 2016-18 | 12.4 (11.4, 13.3) | 7.1 (6.0, 8.4) | 7.9 (7.4, 8.5) |
| | 2017-19 | 11.8 (10.9, 12.7) | 7.1 (5.9, 8.4) | 7.8 (7.2, 8.3) |
| Female | 2000-02 | 8.2 (7.4, 9.0) | 5.4 (4.4, 6.5) | 6.6 (6.1, 7.1) |
| | 2001-03 | 8.4 (7.5, 9.3) | 5.5 (4.5, 6.5) | 6.3 (5.8, 6.8) |
| | 2002-04 | 7.9 (7.0, 8.9) | 5.3 (4.4, 6.3) | 6.4 (5.9, 6.9) |
| | 2003-05 | 7.8 (7.0, 8.8) | 6.2 (5.2, 7.1) | 6.3 (5.9, 6.9) |
| | 2004-06 | 8.4 (7.5, 9.3) | 6.3 (5.4, 7.2) | 6.8 (6.3, 7.2) |
| | 2005-07 | 8.5 (7.4, 9.4) | 6.9 (5.9, 7.8) | 6.6 (6.2, 7.0) |
| | 2006-08 | 8.6 (7.7, 9.5) | 6.6 (5.6, 7.5) | 6.9 (6.4, 7.3) |
| | 2007-09 | 8.3 (7.4, 9.3) | 6.6 (5.6, 7.6) | 6.5 (6.1, 7.0) |
| | 2008-10 | 8.4 (7.5, 9.3) | 5.9 (5.0, 6.8) | 6.6 (6.2, 7.1) |
| | 2009-11 | 8.6 (7.7, 9.6) | 6.5 (5.5, 7.6) | 6.6 (6.1, 7.0) |
| | 2010-12 | 8.4 (7.4, 9.2) | 6.9 (5.8, 7.9) | 6.6 (6.1, 7.1) |
| | 2011-13 | 8.6 (7.7, 9.6) | 7.4 (6.3, 8.4) | 6.6 (6.1, 7.0) |
| | 2012-14 | 8.8 (7.9, 9.6) | 7.3 (6.3, 8.3) | 6.5 (6.0, 6.9) |
| | 2013-15 | 9.0 (8.0, 9.9) | 7.4 (6.4, 8.4) | 6.6 (6.2, 7.0) |
| | 2014-16 | 9.2 (8.2, 10.0) | 7.5 (6.4, 8.5) | 6.8 (6.3, 7.2) |
| | 2015-17 | 9.5 (8.5, 10.5) | 7.2 (6.1, 8.2) | 6.7 (6.2, 7.1) |
| | 2016-18 | 10.0 (9.0, 10.9) | 6.7 (5.7, 7.6) | 6.7 (6.2, 7.2) |
| | 2017-19 | 11.1 (10.0, 12.1) | 6.5 (5.6, 7.4) | 6.5 (6.1, 7.0) |
